## supplementary information for "Fundamental limits on inferring epidemic resurgence in real time using effective reproduction numbers"

### S1 Appendix

#### Methods

We derive some of the mathematical formulae central to the main text. **Eq. (1)** describes the renewal model [1], which simulates the spread of an epidemic, characterising how incidence at some time $s$, $I_s$, depends on the effective reproduction number at that time, $R_s$, and the total infectiousness, $\Lambda_s$. Inference under this model commonly assumes that an incidence window of size $m$ defined as $\tau(s) = \{s, s-1, \dots, s-m+1\}$ contains all the information about $R_s$ [2]. Consequently, we have the Poisson joint log-likelihood over this window, $l_s$, (see Supplement of [3]), with grouped sums $i_{\tau(s)} = \sum_{u \in \tau(s)} I_u$ and $\lambda_{\tau(s)} = \sum_{u \in \tau(s)} \Lambda_u$, as follows.

$$l_s = \log \mathrm{P}(I_{s-m+1}^{s} |\, R_s) = \; i_{\tau(s)} \log R_s - R_s \lambda_{\tau(s)} + \; \zeta_{\tau(s)}. \quad \text{(A)}$$

In **Eq. (A)**, $\zeta_{\tau(s)} = \sum_{u \in \tau(s)} -I_u! + I_u \log \Lambda_u$ is independent of $R_s$. The maximum likelihood $\tilde{R}_s$ estimate under this model solves $\frac{\partial l_s}{\partial R_s} = 0$ i.e., $\tilde{R}_s = i_{\tau(s)} \lambda_{\tau(s)}^{-1}$. The Fisher information, $\mathrm{FI}[R_s]$, defines the best achievable precision (i.e., smallest variance) around this estimate [4], and is computed from **Eq. (A)** as $\mathrm{E}\left[-\frac{\partial^2 l_s}{\partial R_s^2}\right]$ [3,4]. This gives $\frac{\mathrm{E}[\sum_{u \in \tau(s)} I_u]}{R_s^2}$. Substituting $\mathrm{E}[I_u] = \Lambda_u R_s$ from **Eq. (1)**, then yields the key result in the left side of **Eq. (2)** in the main text.

Widely used real-time methods, such as *EpiEstim* [2] and related approaches, often calculate the posterior distribution $\mathrm{P}(R_s | I_1^s) \approx \mathrm{P}(R_s | I_{s-m+1}^s)$. This approximation is a consequence of the *m*-window assumption and is conventionally obtained by setting a conjugate gamma prior distribution i.e., $\mathrm{P}(R_s) \equiv \mathrm{Gam}(a, \; c^{-1})$. Hyperparameters (e.g., $a = 1, \; c = 1/5$) are often selected to ensure this prior distribution is uninformative. Applying Bayes law with the Poisson likelihood from **Eq. (A)** yields $\mathrm{P}(R_s | I_{s-m+1}^s) \equiv \mathrm{Gam}\left(a + i_{\tau(s)}, \; \left(c + \lambda_{\tau(s)}\right)^{-1}\right)$.

We can compute the resurgence probability as $\mathrm{P}(R_s > 1 |\, I_1^s) \approx \int_1^\infty \mathrm{P}(R_s | I_{s-m+1}^s)\, dR_s$. This approximation also proceeds from the window-based formulation. The cumulative distribution function of the gamma posterior distribution, $\mathrm{F}_s(x)$, can be written as below for some $x$.

$$\mathrm{F}_s(x) = \mathrm{P}(R_s \leq x | I_{s-m+1}^{s}) = 1 - \sum_{j=0}^{a-1+i_{\tau(s)}} \frac{x^j \left(c + \lambda_{\tau(s)}\right)^j}{j!} e^{-x\left(c+\lambda_{\tau(s)}\right)}. \quad \text{(B)}$$

**Eq. (B)** results from standard properties of gamma distributions. We compute the resurgence probability as $1 - \mathrm{F}_s(1)$, which gives the right side of **Eq. (2)** in the text. The above formulae are useful both for providing analytic insight and measuring performance of realistic estimators used in outbreak analysis, which adhere to this formulation [2,5–7].

These equations all feature a dependence on the choice of window size *m*. As investigated in [3] large *m* can mean that we are slower to detect transmissibility changes, while small *m* can lead to oversensitivity to noise. We avoid this *m*-dependence by simply using this approach to gain general, theoretical insights into detection asymmetries and latencies. Specifically, in the main text we prove that the lag in inferring resurgence is larger than that when estimating a corresponding control signal, for arbitrary window sizes (due to smaller historical incidence across suspected periods of resurgence). We then perform more detailed (but less tractable) investigations to discern the likely magnitude of these asymmetric lags.

These investigations (in **Figs 2-3** of the main text and **Fig A** below) apply the *EpiFilter* method [8], which circumvents window size issues. *EpiFilter* exploits formal signal processing theory to minimise the mean squared error in the estimation of $R_s$. Its sequential predictive accuracy (i.e., it has small generalisation error) and its ability to detect change-points in real time have been verified on extensive simulations [8,9], suggesting it as a largely assumption-free tool, suitable for exploring fundamental limits on resurgence and control. This difference in methodology is signified in our notation in **Eq. (3)** of the text, which no longer uses window approximations ($\tau(s)$). There our results are direct outputs of *EpiFilter*.

Derivations for the inference equations behind the filtering and smoothing in *EpiFilter* are in [8,10]. This more general formulation allows us to go beyond the analytic insights from the *EpiEstim* type models above and limits the influence of prior distributions on results (which is particularly strong when incidence is small) since $R_s$ is a-priori uniformly distributed over some wide range ([0.01, 10] here). Consequently, we examine the problem of resurgence detection from multiple angles. The prior distributions used in all methods have mean and median above 1 so that any delays we find in detecting resurgence are the absolute minimum possible.

The trends uncovered in **Eq. (4)** of the main text, where heterogeneity is explored, are within the *EpiEstim* framework, but will be valid for *EpiFilter* and general *R*-estimation methods, since they result from the properties of convex sums and averages only. Last, while our conclusions may appear limited due to their dependence on renewal models, we note that renewal models (i) can describe realistic transmission patterns for many diseases with accuracies comparable to that of more detailed network-based models [11] (ii) are the dominant model for measuring real-time outbreak changes [7,11,12] and (iii) are able to equivalently represent the dynamics of prevailing compartmental models, such as the SEIR model, depending on the form of the generation time distribution considered [13].

### Additional Figures

We provide simulations in **Fig A** for simulated COVID-19 epidemics, showing that significant delays in detecting resurgence but not epidemic control persist. These are consistent with and support **Fig 2** of the main text, which examined Ebola virus disease dynamics. While figures plot ensembles of mean estimates, single simulations (where the estimated credible intervals reflect noise from the incidence of that simulation) also display this asymmetry, confirming that real-time resurgence detection is innately hard.

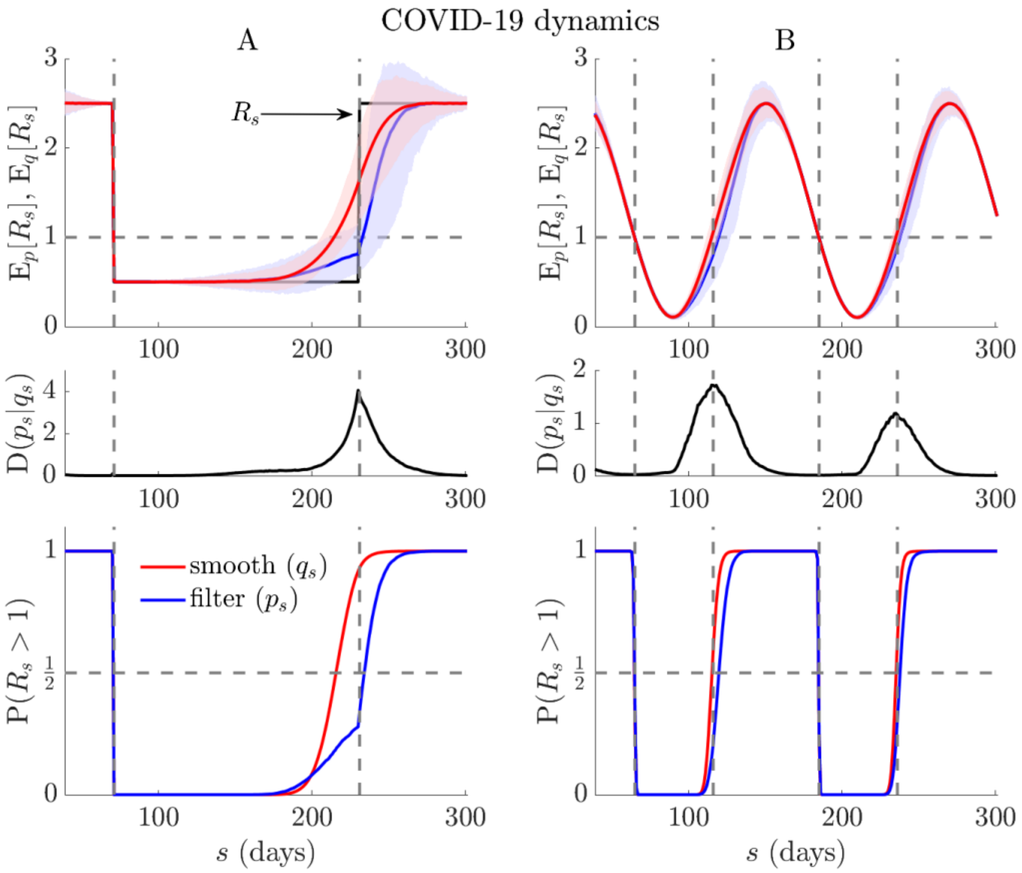


**Fig A: Resurgence and control dynamics of COVID-19.** We repeat the simulations from **Fig 2** (main text) but for 1000 realisations of COVID-19 epidemics ($t = 300$) using generation times from [14] (true $R_s$ in black). Top panels show the posterior mean estimates from every realisation (computed via *EpiFilter* [8]). Middle panels average Kullback-Liebler divergences from those simulations, $D(p_s|q_s)$, and bottom panels show the overall filtered ($P(R_s > 1 \mid I_1^s)$, blue), and smoothed ($P(R_s > 1 \mid I_1^t)$, red) probabilities of resurgence. In keeping with **Fig 2**, we find fundamental and appreciable latencies in detecting resurgence, often an order of magnitude longer than those for detecting epidemic control (compare red and blue curves in relevant panels). The initial rise in $P(R_s > 1 \mid I_1^s)$ of panel A, which precedes the $R_s$ change is due to the prior distribution over $R_s$ (which has mean > 1) in a period with very few cases.

We also provide plots of the simulated incidence curves (i.e., daily counts of infected cases), which underlie the results of **Fig A** above and **Fig 2** of the main text, in **Fig B** and **Fig C** respectively. The generally smaller incidence (which is also stochastically noisier) that often associates with resurgence events contributes to their innate detection difficulty.

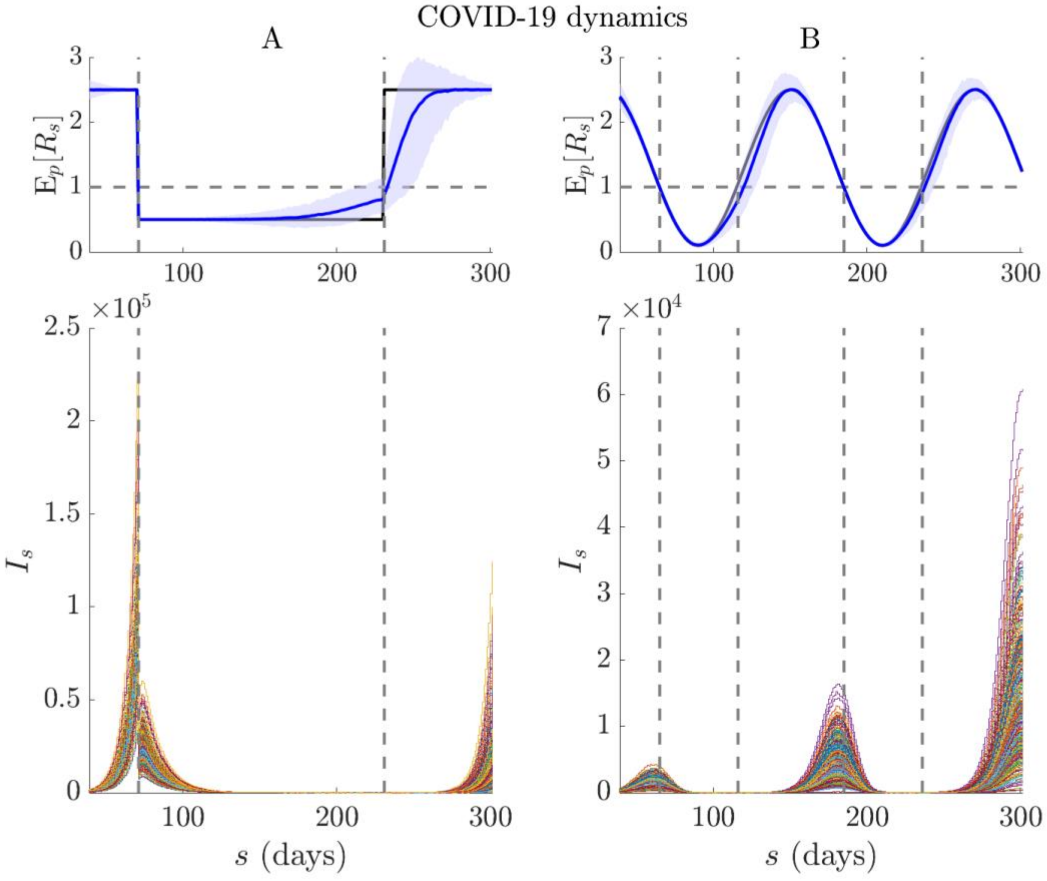


**Fig B: Incidence curves for COVID-19.** We present the counts of daily new cases, $I_s$, (bottom panels), which are simulated from the $R_s$ trajectories (black, top panels) depicting a rapid control and resurgence (A) and a seasonally varying transmissibility (B). These curves are generated using renewal models (see Methods above) with COVID-19 generation times from [14] and underlie the results in **Fig A**. We include the filtered (real-time) estimates, $E_p[R_s]$, from that figure for completeness (blue with 95% credible intervals).

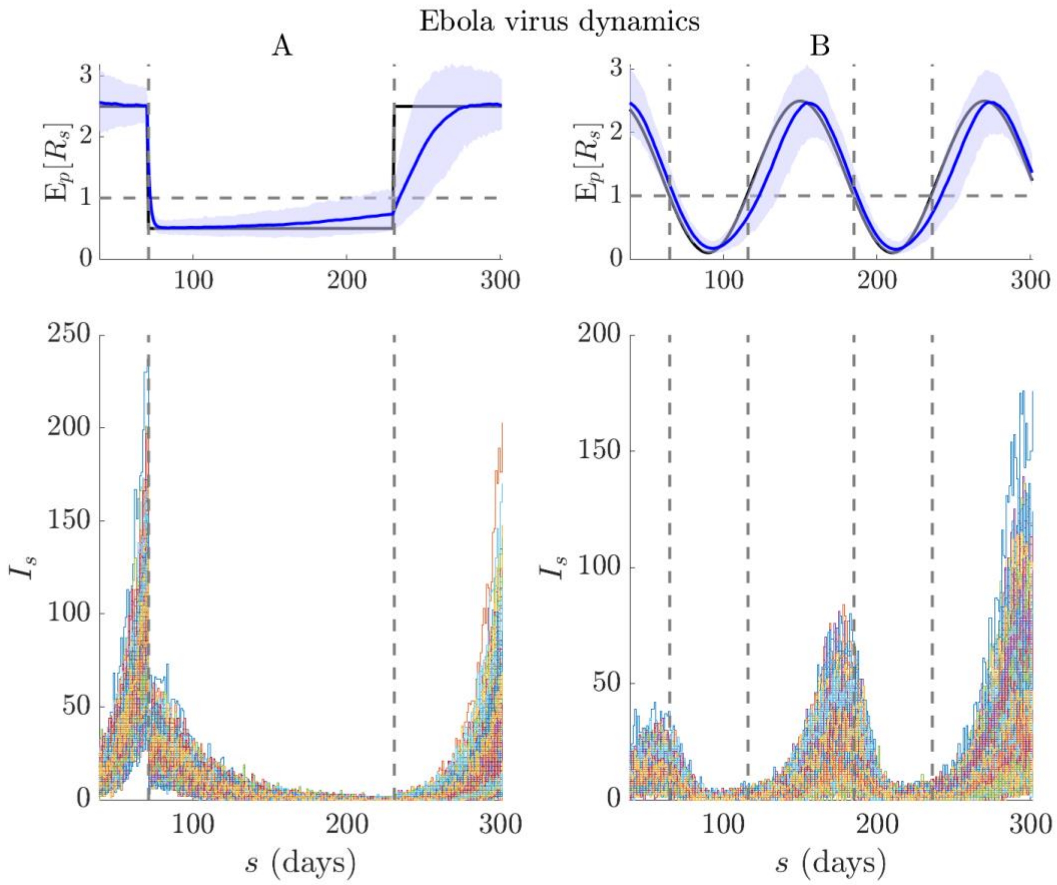


**Fig C: Incidence curves for Ebola virus disease.** We present the counts of daily new cases, $I_s$, (bottom panels), which are simulated from the $R_s$ trajectories (black, top panels) depicting a rapid control and resurgence (A) and a seasonally varying transmissibility (B). These curves are generated using renewal models (see Methods above) with Ebola virus generation times from [15] and underlie the results in **Fig 2** of the main text. We include the filtered (real-time) estimates, $E_p[R_s]$, from that figure for completeness (blue with 95% credible intervals).
